# Protocol for Development of a Reporting Guideline for Causal and Counterfactual Prediction Models

**DOI:** 10.1101/2021.11.19.21266604

**Authors:** Jie Xu, Yi Guo, Fei Wang, Hua Xu, Robert Lucero, Jiang Bian, Mattia Prosperi

## Abstract

**Introduction:** While there are protocols for reporting on observational studies (e.g., STROBE, RECORD), estimation of causal effects from both observational data and randomized experiments (e.g., AGREMA, CONSORT), and on prediction modelling (e.g., TRIPOD), none is purposely made for assessing the ability and reliability of models to predict counterfactuals for individuals upon one or more possible interventions, on the basis of given (or inferred) causal structures. This paper describes methods and processes that will be used to develop a reporting guideline for causal and counterfactual prediction models (tentative acronym: PRECOG).

**Materials and Methods:** PRECOG will be developed following published guidance from the EQUATOR network, and will comprise five stages. Stage 1 will be bi-weekly meetings of a working group with external advisors (active until stage 5). Stage 2 will comprise a scoping/systematic review of literature on counterfactual prediction modelling for biomedical sciences (registered in PROSPERO). In stage 3, we will perform a computer-based, real-time Delphi survey to consolidate the PRECOG checklist, involving experts in causal inference, statistics, machine learning, prediction modelling and protocols/standards. Stage 4 will involve the write-up of the PRECOG guideline (including its checklist) based on the results from the prior stages. In stage 5, we will work on the publication of the guideline and of the scoping/systematic review as peer-reviewed, open-access papers, and on their dissemination through conferences, websites, and social media.

**Conclusions:** PRECOG can help researchers and policymakers to carry out and critically appraise causal and counterfactual prediction model studies. PRECOG will also be useful for designing interventions, and we anticipate further expansion of the guideline for specific areas, e.g., pharmaceutical interventions.

## 1 BACKGROUND

The increasing availability of large electronic health record data has led to an explosion in the development of prediction models –both traditional statistics and machine learning– for diagnostic, prognostic, and treatment optimization purposes. Despite of the availability of reporting guidelines, e.g., “transparent reporting of a multivariable prediction model for individual prognosis or diagnosis” (TRIPOD)^1^, the quality of many studies is low, as well as adherence to reporting standards, and there is often misinterpretation of the models’ operating capabilities, with possible misuse and harm at the individual and/or population level^2,3^. One of the most common mistakes is to consider a prediction model readily usable for interventions on individuals, by changing certain variables with the intent to improve outcomes, i.e., calculating alternative scenarios or so-called counterfactuals. Since prediction models are often learnt from observational data, there is no guarantee that the strongest predictors are causing the outcome of interest and are not confounded, mediated by others, or actually concomitant causes of it. While such bias is not a problem for mere prediction in similar populations –since variables are not being changed with the intent to modify risk– it becomes problematic on new populations (even with high cross-validation results)^4^ and when trying to optimize outcomes^5^.

Thus, formal causal assessment is needed when developing prediction models on observational data to be used for alternative scenarios and interventions, i.e., counterfactual prediction models. The approaches from traditional statistics, computational science, and econometrics, including the potential outcomes framework^6^, do-calculus and directed acyclic graphs (DAGs)^7^, are often focused on estimating a population-level causal effect for a single interventional query (treatment or exposure), but in principle can be used to calculate individual treatment effects and counterfactuals. Machine learning has also been employed for counterfactual prediction^8,9^. Several off-the-shelf methodologies have been revisited, including deep learning^10–13^, and random forests^14^.

Given the rise in counterfactual prediction modelling studies, there is need for common grounds on model reporting, to improve on overall quality (albeit adhering to a protocol might be necessary, yet not sufficient condition to study quality), and specifically on transparency and reproducibility of results.

In the “Enhancing the quality and transparency of health research” (EQUATOR) network (https://www.equator-network.org/), there are guidelines specifically designed for reporting causal effects on RCTs, e.g., “consolidated standards of reporting trials” (CONSORT)^15^ and “a guideline for reporting mediation analyses of randomized trials and observational studies” (AGREMA)^16^. Reporting guidelines for observational studies also mention causal effects inference, e.g., “strengthening the reporting of observational studies in epidemiology Using Mendelian randomization” (STROBE-MR)^17^, “reporting of studies conducted using observational routinely collected health data statement for pharmacoepidemiology” (RECORD-PE)^18^, and the “instrumental variable methods in comparative safety and effectiveness research”^19^. Outside of EQUATOR, the Patient-Centered Outcomes Research Institute (PCORI) (https://www.pcori.org/) provides “Standards for Causal Inference Methods in Analyses of Data from Observational and Experimental Studies in Patient-Centered Outcomes Re-search” (https://tinyurl.com/4x55ad3t). Also, there are guidelines for for estimating causal effects in pragmatic randomized trials^20^.

Overall, existing guidelines are not well fitted for causal and counterfactual prediction modelling, although a number of them contain elements that are directly related. Consequently, we aim to develop a new reporting guideline, which we tentatively name as “prediction and counterfactual modelling guidelines” (PRECOG). The focus of PRECOG is the development and validation of counterfactual prediction models, where one or more variables can be intervened upon, and will require declaration of causal assumptions as well validation of causal claims. PRECOG will also cover software implementation and interoperability. The primary use cases of PRECOG are expected to fall within biomedical sciences, but they could be applied to other fields such as psychology or economics.

## 2 METHODS

PRECOG will be developed following published guidance from the EQUATOR network^21^. We will develop the guideline in five stages: (1) bi-weekly meeting of a working group; (2) scoping/systematic review of causal and counterfactual prediction modelling studies; (3) reporting checklist draft and Delphi exercise; (4) development of the final guideline; and (5) peer-review, publication and dissemination. These stages are drawn from prior, successful development studies, in primis the protocol used for the making of TRIPOD-AI and PROBAST-AI^22^.

**Figure 1.**
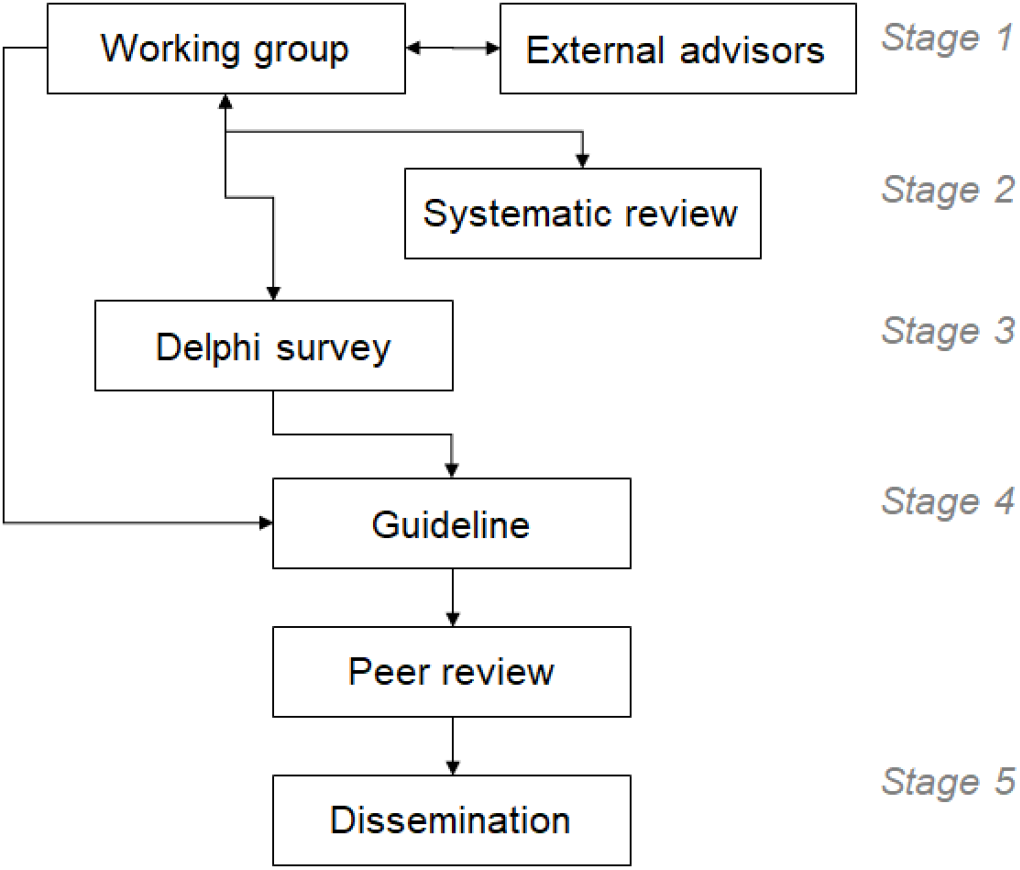
Flowchart of the PREdiction and COunterfactual modelling Guidelines (PRECOG) development.

### 2.1 Stage 1: Working Group Setup and Meetings

The core working group is composed by the co-authors of this protocol, who met bi-weekly (30-45 minutes) since September 13, 2021 to discuss the development of the reporting guideline. After the public posting of the protocol, the working group will be expanded with external advisors with expertise in biomedical informatics, (bio)statistics, causal inference, computer science, epidemiology, health economics, health outcome research, standards, and related areas. Each member of the core working group will identify one or more suitable external advisors, who will be invited to participate the meeting and prompted to suggest further advisors. The list of advisors will be also be used for Stage 3 (Delphi exercise). The working group will make best efforts to assure diversity, variety in career stages, and multicultural representation. The extended working group will meet also bi-weekly, and each meeting will ideally be composed by 3-7 people, with at least one external advisor present (otherwise be rescheduled). The working group will work on: (a) review of existing EQUATOR/PCORI reporting guidelines; (b) evaluation of the results of the scoping/systematic review of counterfactual prediction modelling studies for biomedical sciences; (c) drafting of the initial reporting checklist for the Delphi survey; (d) review of the survey and development of the final guideline; (e) manuscript writing; and (f) submission of the products to peer-review, publication and dissemination.

### 2.2 Stage 2: Literature Review of Counterfactual Prediction Modelling Studies

The purpose of the literature review is twofold: (1) to build a knowledge base on study design, methodological approaches, use cases and reporting commonalities among causal inference and counterfactual prediction studies in biomedical sciences; and (2) to help development of reporting items for PRECOG. A subset of the working group members will concentrate on the review. After determining the overarching objective, search criteria and performing an initial screening, the team will decide if a scoping review will be preferred to a systematic review^23^. The planned reporting statement of choice is the “preferred reporting items for systematic reviews and meta-analyses” (PRISMA)^24^, which includes also an extension for scoping reviews, and the working group will register the work in the “prospective register of systematic reviews” (PROSPERO)^25^.

### 2.3 Stage 3: Delphi Exercise

We will conduct a Delphi survey to review and refine the items of the PRECOG reporting checklist. Delphi participants will be identified initially through the professional network of the core working group and of the external advisors, and further via literature search (including but not limited to the scoping/systematic review), social media screening, and snowballing by the active participants. As for the expanded working group composition, participants will be invited from diverse and multicultural background and different countries. Invitees will include academics at various career stages, researchers and investigators from non-profit and for-profit organizations, program officers from national/federal funding agencies, entrepreneurs, health care professionals, journal editors, policy makers, health care regulators, and end-users of predictive models. The working group will also discuss and agree on a suitable sample size for the Delphi survey.

We will employ computer-based, real-time Delphi, which offers some operational advantages with respect to traditional multi-round Delphi techniques^26^. The working group will develop an initial reporting checklist for PRECOG, based on the EQUATOR developing standard, existing related guidelines (e.g., TRIPOD, PCORI), and an anonymous online survey will be created where each checklist item can be evaluated in relation to its importance and relevance for the guideline, using a five-point Likert scale, and a free text box for comments. Also, at the end of the survey, another text box will allow more generic comments and propositions, e.g., new items to be added to the checklist. When a participant consents to participate and completes the survey for the first time, they receive a summary of all the responses to date, and a code to access the survey again within the next three weeks. Each participant can see the updated results within that time frame and make changes to their responses if they deem so. The survey is closed after the required sample size is reached, or a maximum of two months are passed from the first recorded response.

At the end of the Delphi survey, the working group will review the results and consolidate the checklist. Items will need to reach 80% agreement from the panel in order to be accepted (or omitted) in the development of the final guideline. Eighty percent was chosen as an appropriate cut off based on work by Lynn^27^, who suggested that when at least 10 experts are involved in consensus development, at least 80% of the experts must agree on an item to achieve content validity. Statements that do not meet the 80% agreement will be discussed during the bi-weekly meetings, and dropped if no consensus is reached by the extended working group.

### 2.4 Stage 4: Development of the Guideline and Related Products

Upon finalization of the reporting checklist from the Delphi exercise, the extended working group will develop the full PRECOG guideline. The manuscript will be posted to a public pre-print website, e.g., bioRxiv or medRxiv, before submission to a peer-review journal, and possibly presented as abstract/poster in major international conferences, e.g, the annual conference of the American Medical Informatics Association (AMIA) or the Society for Epidemiology Research (SER). It is expected that the PRECOG initiative will produce at least the following papers:

- Guideline development protocol (this work);
- Scoping/systematic review or causal and counterfactual prediction models in biomedical sciences;
- PRECOG guideline.

### 2.5 Stage 5: Publication and Dissemination Plan

After being posted on pre-print servers, the aforementioned manuscripts will be submitted to peer-reviewed international journals for final publication. The authors’ list will be determined on the basis of effective individual contributions, following the “contributor roles taxonomy” (CRediT) (https://casrai.org/credit/), and might include additional contributors other than the working group members and external advisors.

The dissemination strategy will be discussed during the bi-weekly meetings. In addition to conferences and publications, it is likely that social media platforms such as Twitter will be leveraged to inform on the PRECOG availability and utility.

## 3 CONCLUSION

The number of causal inference and counterfactual prediction modelling studies, along with software development, is increasing rapidly. PRECOG can help researchers and policymakers to carry out and critically appraise these studies and tools, besides providing model developers with a transparent and reproducible framework, and liaising with model updating and evidence synthesis projects. PRECOG will also be useful for designing interventions, and we anticipate further expansion of the guideline for specific areas, e.g., pharmaceutical interventions. The guideline will be periodically reviewed to ensure consistency with the EQUATOR standards and with best methodological, operational scientific, and ethical practices.

## Data Availability

N/A

## 4 ACKNOWLEDGMENTS

This work has been in part supported by National Institutes of Health (NIH) - National Institute of Allergy and Infectious Diseases (NIAID) grants no. R01AI145552 and R01AI141810 (Dr. Prosperi), by National Institute on Aging (NIA) grants no. R33AG062884-03 (Dr. Lucero and Dr. Prosperi) and 5R21AG068717-02 (Dr. Bian and Dr. Guo), by National Cancer Institute (NCI) grants no. 5R01CA246418-02, 3R01CA246418-02S1, 1R21CA245858-01A1, 3R21CA245858-01A1S1, and 1R21CA253394-01A1 (Dr. Bian and Dr. Guo), and by Centers for Disease Control and Prevention (CDC) grant no. U18DP006512 (Dr. Bian, Dr. Guo and Dr. Prosperi).

